# Dynamic Detection of Delayed Cerebral Ischemia Using Machine Learning

**DOI:** 10.1101/2020.04.15.20067041

**Authors:** Murad Megjhani, Kalijah Terilli, Ayham Alkhachroum, David J. Roh, Sachin Agarwal, E. Sander Connolly, Angela Velazquez, Amelia Boehme, Jan Claassen, Soojin Park

**Author notes:** **Corresponding Author:** Soojin Park, 177 Fort Washington Ave, 8 Milstein-300Center, NY, NY 10032. contributed equally (Murad Megjhani, Kalijah Terilli). **Potential Conflicts-of-Interest/Disclosures:** Funding: This study was funded by National Institute of Health (NIH), grant number: NIEHS K01- ES026833-02 (SP) and American Heart Association grant number: 20POST35210653 (MM). **Compliance with Ethical Standards:** Ethics approval and consent to participate: The study was approved by the Columbia University Medical Center Institutional Review Board. In all cases, written informed consent was obtained from the patient or a surrogate. **Statistical Analysis:** conducted by Dr. Amelia Boehme and Dr. Murad Megjhani. **Study Funded:** by NIEHS K01-ES026833-02 (SP). **Authors’ contributions:** Data Collection (KT, MM, SP, AV, DR, SA, JC), Analysis (MM, KT, SP, AB), Writing (KT, MM, SP, AA), Editing (All).

## Abstract

**Objective:** To develop a machine learning based tool, using routine vital signs, to assess delayed cerebral ischemia (DCI) risk over time.

**Methods:** In this retrospective analysis, physiologic data for 540 consecutive acute subarachnoid hemorrhage patients were collected and annotated as part of a prospective observational cohort study between May 2006 and December 2014. Patients were excluded if (i) no physiologic data was available, (ii) they expired prior to the DCI onset window (< post bleed day 3) or (iii) early angiographic vasospasm was detected on admitting angiogram. DCI was prospectively labeled by consensus of treating physicians. Occurrence of DCI was classified using various machine learning approaches including logistic regression, random forest, support vector machine (linear and kernel), and an ensemble classifier, trained on vitals and subject characteristic features. Hourly risk scores were generated as the posterior probability at time *t*. We performed five-fold nested cross validation to tune the model parameters and to report the accuracy. All classifiers were evaluated for good discrimination using the area under the receiver operating characteristic curve (AU-ROC) and confusion matrices.

**Results:** Of 310 patients included in our final analysis, 101 (32.6%) patients developed DCI. We achieved maximal classification of 0.81 [0.75-0.82] AU-ROC. We also predicted 74.7 % of all DCI events 12 hours before typical clinical detection with a ratio of 3 true alerts for every 2 false alerts.

**Conclusion:** A data-driven machine learning based detection tool offered hourly assessments of DCI risk and incorporated new physiologic information over time.

## Introduction

Delayed cerebral ischemia (DCI) is heavily linked with poor outcomes after aneurysmal subarachnoid hemorrhage (aSAH), and occurs in 21-33% of patients.^1-3^ DCI has been implicated as the most morbid secondary injury after aSAH, showing independent association with cognitive impairment.^4^ Despite the high impact on patient outcomes, there are many barriers to timely detection of DCI. Symptom onset can be subtle and develop gradually, and clinical diagnosis often relies on a change in exam, though 20% of DCI patients are asymptomatic.^2^ Physicians must surpass a certain threshold of suspicion to warrant the health risks of confirmatory testing (radiation exposure, laying patient flat), which includes ruling out clinical mimics.^5^ There is an opportunity for new technology to help improve the timely diagnosis of DCI.

Transcranial doppler is the only non-invasive surveillance tool supported by guidelines, yet it is limited by poor sensitivity, poor inter-rater reliability and technician availability, also adequate insonation is not possible for 10-15% of patients.^6,7^ Continuous quantitative electroencephalogram (qEEG) has shown promising results for detecting DCI, however implementation requires 24 hour continuous EEG monitoring and clinical expert artifact reconciliation and event labeling.^8,9^ An optimal DCI monitoring tool would be continuous and automated, performing without reliance on expert or technician availability.

In prior work, we featurized physiologic signals (heart rate, blood pressure, oxygen saturation) for the classification of DCI patients, reporting good prediction results (AUROC 0.78) that surpassed modified Fisher Scale alone (AUROC 0.54-0.58).^10,11^ While the success of these models supports the existence of a useful physiologic signal for DCI classification and improves precision of prediction for individuals, they incorporated information only from the first few days of admission and offer a static, one-time assessment of risk. With such a design, no insight can be offered regarding when DCI will develop within the large window of onset (3-21 days post-bleed). In the present study, we apply machine learning methods to develop algorithms utilizing routine, continuous physiologic information to detect when DCI becomes more or less likely to occur.

Machine learning derived detection tools have been successfully developed and implemented in other areas of critical care, including but certainly not limited to: weaning in mechanically ventilated patients^12^, instability and risk of cardiac arrest in children^13^, hemodynamic shock ^14^, acute kidney injury ^15,16^ and extensively in sepsis^17-20^. We propose a data-driven DCI detection tool that offers hourly estimations of the patient’s current state, incorporating new physiologic information over time. If proven successful, such a model could be used to establish DCI onset time for better evaluation of proposed treatments and to improve patient care by acting as a trigger for confirmatory testing and subsequent timely intervention.

## Methods

### Study Population

Patients with aneurysmal SAH consecutively admitted to the neurological intensive care unit (NICU) were prospectively enrolled in an observational cohort study which has been previously been described.^21^ Baseline demographics used for this study are *core* and *supplemental-highly recommended* National Institute of Neurological Disorders and Stroke (NINDS) common data elements (CDEs) for SAH subject characteristics: age, gender, ethnicity, tobacco use, hypertension history and premorbid modified Rankin Scale (mRS). Routinely collected grading scales of injury severity and outcome prediction were the *core* CDE: World Federation of Neurological Surgeons scale (WFNS), and the *supplemental* CDEs: modified Fisher Score (mFS), Hunt & Hess grade (HH) and Glasgow Coma Scale (GCS).^22,23^ Comorbidities taken into account for this study were cerebral edema, fever (>38.3°C), pulmonary edema, hydrocephalus, and seizure. Outcome variables were in-hospital mortality, length of stay in the NICU and modified Rankin Scale (mRS) at discharge, 3 months and 12 months post-discharge.

The target classification outcome of the study was DCI, which was determined by consensus of attending physicians when patients met the following criteria: delayed neurological deterioration defined as a ≥ 2 point change in GCS or new focal neurological deficit lasting for > 1 hour and not associated with surgical treatment, and/or a new cerebral infarct on brain imaging which is not attributable to any other causes.^24-26^ Clinical management of SAH patients adhered to the Neurocritical Care Society and American Heart Association guidelines (**Appendix A**).^1,16^ All patients from this prospectively collected and annotated cohort were included in this retrospective analysis, unless they met one of the following exclusion criteria: no parametric physiologic data was available, they expired prior to the DCI onset window (< PBD 3), or early angiographic vasospasm was detected on admitting angiogram (as DCI development was the signal of interest).

### Physiologic Data

Vitals data was collected using a high-resolution acquisition system (BedmasterEX; Excel Medical Electronics Inc, Jupiter, FL, USA) from General Electric Solar 8000i monitors (Port Washington, NY, USA; 2006-2014) at 0.2 Hz. Heart rate (HR), intracranial pressure (ICP), respiratory rate (RR), oxygen saturation (SPO2), systolic and diastolic arterial blood pressure (SBP, DBP) and temperature (TEMP) were extracted and analyzed using custom software developed in Matlab 2016a (Mathworks, Natick, MA).

The large window for DCI presentation makes direct comparison across patients over time difficult. To address this, we use DCI diagnosis as the temporal anchor to align the data so that we can identify a physiologic signal as the disease develops, despite varied times of onset (**Appendix B**). Data for each physiologic measure was first rescaled to allow for comparison using min-max normalization.^27^ There were 20 total time-varying measures computed in 10-minute windows: mean and standard deviation (SD) for each of the seven vitals, and maximum cross-correlation (lag –30 to +30 seconds) of HR with each remaining six vital sign (ICP, RR, SPO2, SBP, DBP and TEMP). Summary statistics were only retained for segments with at least 50% of the data available. Finally, hourly averages were obtained, resulting in 336 time points over 14 days. Analysis was performed in Matlab 2016a (Mathworks, Natick, MA).

### Statistical analysis

#### Comparison of Demographics

Demographic data were summarized and compared for the DCI and control groups. Fisher’s exact test was applied to categorical variables, and the Mann-Whitney U test for two-group comparisons was applied to continuous variables. All statistical tests were two-tailed and *p*-value < 0.05 was considered statistically significant. Analysis was performed in R Studio software (version 1.0.143, http://www.rstudio.com, RStudio Inc., Boston, USA).

#### Comparison of Vital Signs

Multilevel linear regression was used to identify points of statistically significant difference between DCI+ and DCI– time series data. Windowing for each comparison was done using a cumulative strategy to mimic a clinically-relevant setting, so in the future information could be added over time as a prospective patient would continuously generate new data, see **Appendix C**. We employed a repeated-measures design in this study due to the repeated outcomes. Since the physiologic data were collected over time, we used a longitudinal mixed effects model to assess the relationship between DCI and the physiologic time series data. This method enabled us to model intra- and inter-patient variability. We applied the mixed-effects model for each outcome of interest, and assessed how the groups differed for the 14 days surrounding the DCI anchor. For more information about these models, see **Appendix C**. Statistical analysis was conducted using SAS (SAS Institute, Cary, NC).

### Machine Learning

To translate our findings into a tool for classification, we applied an ensemble machine learning approach. We computed the range (max-min), mean, SD, median, IQR and entropy for each of the 20 vital sign measures and added 6 subject characteristic variables (age, sex, mFS, WFNS, HH and GCS at NICU admission), resulting in 126 total features. Missing values were imputed using the median. We used F-statistics to identify *k* number of best features explaining most variance within the dataset ^28^. We then built L2-regularized logistic regression (LR), linear and kernel support vector machine (SL, SK), random forest (RF), and ensemble (EC) classifiers for each day prior to the DCI anchor (**Appendix D**) ^29^. We performed nested five-fold cross-validation to tune model parameters and to report the accuracy. More details of how nested cross-validation is used to prevent over-fitting are provided in **Appendix E**. All classifiers were evaluated for good discrimination using the area under the receiver operating characteristic curve (AU-ROC) and confusion matrix.

### Hourly Risk Scores

We generated hourly risk scores indicating current likelihood of DCI using the ensemble classifier at DCI anchor. Risk scores above the optimal cut off point (threshold) indicate higher probability of the patient developing DCI. Given a patient *i* the risk score at time *t* with features *x*_*it*_ is computed as a posterior probability given by:

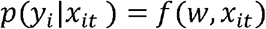

Where *f* is the classifier (EC). Machine learning models and risk scores were developed using the Python scikit-learn library.^30^

## Data availability

All relevant data are presented within the article and its supporting information files. Additional information can be obtained upon request to the corresponding author.

## Results

### Patient Cohort

Clinical and physiological data was available for 540 consecutive SAH patients admitted to the NICU at Columbia University from May 2006 to December 2014. 156 patients met initial exclusion criteria: 27 patients showed vasospasm (VSP) on vessel imaging at NICU admission, aneurysm was not confirmed by angiogram for 98 patients and 31 patients died before entering the DCI window (before PBD 3). In addition, 74 patients displayed angiographic VSP without development of DCI and were removed from the final analysis (**Figure 1**). 310 patients were included in the final analysis for this study; 101 patients developed DCI while 209 patients did not develop DCI and were used as controls (DCI+ 32.6%, DCI– 67.4%). For DCI+ patients the mean PBD of the anchors was 7 ± 3 days (compared to 7 ± 0 days for the DCI– anchors), therefore on average the window used for analysis for all patients was the first 14 days post-bleed. Demographic statistical differences between groups are reported in **Table 1**.

**Table 1.** Characteristics of SAH patients with and without DCI.

|  | DCI+<br>n=101 | DCI-<br>n=209 | *p value* * |
| --- | --- | --- | --- |
| Age, y, median (IQR) | 50 (44-62) | 56 (47-66) | **0.033** |
| Female Gender, n (%) | 81 (80.2) | 137 (65.6) | **0.008** |
| Hispanic Ethnicity, n (%) | 30 (29.7) | 69 (33.0) | 0.604 |
| Tobacco Use, n (%) | 55 (54.5) | 111 (53.1) | 0.713 |
| Hypertension History | 39 (38.6) | 107 (51.2) | **0.040** |
| Pre-Morbid mRS, 3 - 6, n (%) | 3 (3.0) | 7 (3.3) | 1 |
| HH, 4 - 5, n (%) | 40 (39.6) | 48 (23.0) | **0.003** |
| WFNS, 4 - 5, n (%) | 50 (49.5) | 71 (34.0) | **0.009** |
| GCS, median (IQR) | 13 (6-15) | 15 (9-15) | **<0.001** |
| mFS, 3 - 4, n (%) | 38 (37.6) | 83 (39.2) | 0.805 |
| mFS 0, n (%) | 0 (0.0) | 15 (7.2) |  |
| mFS 1, n (%) | 15 (14.9) | 41 (19.6) |  |
| mFS 2, n (%) | 48 (47.5) | 71 (34.0) |  |
| mFS 3, n (%) | 32 (31.7) | 73 (34.9) |  |
| mFS 4, n (%) | 6 (5.9) | 9 (4.3) |  |
| Cerebral Edema, n (%) | 76 (75.2) | 100 (47.8) | **<0.0001** |
| Fever >38.6°C, n (%) | 54 (53.5) | 72 (34.4) | **0.002** |
| Hydrocephalus, n (%) | 61 (60.4) | 82 (39.2) | **<0.001** |
| Pulmonary Edema, n (%) | 27 (26.7) | 24 (11.5) | **0.001** |
| Seizures, n (%) | 12 (11.9) | 20 (9.6) | 0.553 |
| In-Hospital Mortality, n (%) | 8 (7.9) | 26 (12.4) | 0.253 |
| LOS, d, median (IQR) | 22 (17-30) | 12 (10-16) | **<0.0001** |
| mRS at Discharge, 3 - 6, n (%) | 89 (88.1) | 106 (50.7) | **<0.0001** |
| mRS at 3 Months, 3 - 6, n (%) | 40 (39.6) | 52 (24.9) | **0.001** |
| mRS at 12 Months, 3 - 6, n (%) | 36 (35.6) | 57 (27.3) | **0.029** |

\* *p* values were calculated using Fisher's exact test for frequency comparisons of categorical variables and Mann-Whitney *U* test for two-group comparisons of continuous variables. All statistical tests were two-tailed. Significant values (*p* < 0.05) are highlighted in bold.
SAH = subarachnoid hemorrhage, DCI = delayed cerebral ischemia, y = years, SD = standard deviation, n = number of patients, mRS = modified Rankin Scale, HH = Hunt & Hess grade, mFS = modified Fisher Scale, WFNS = World Federation of neurological Surgeons scale, GCS = Glasgow Coma Scale, LOS = length of stay

**Figure 1:**
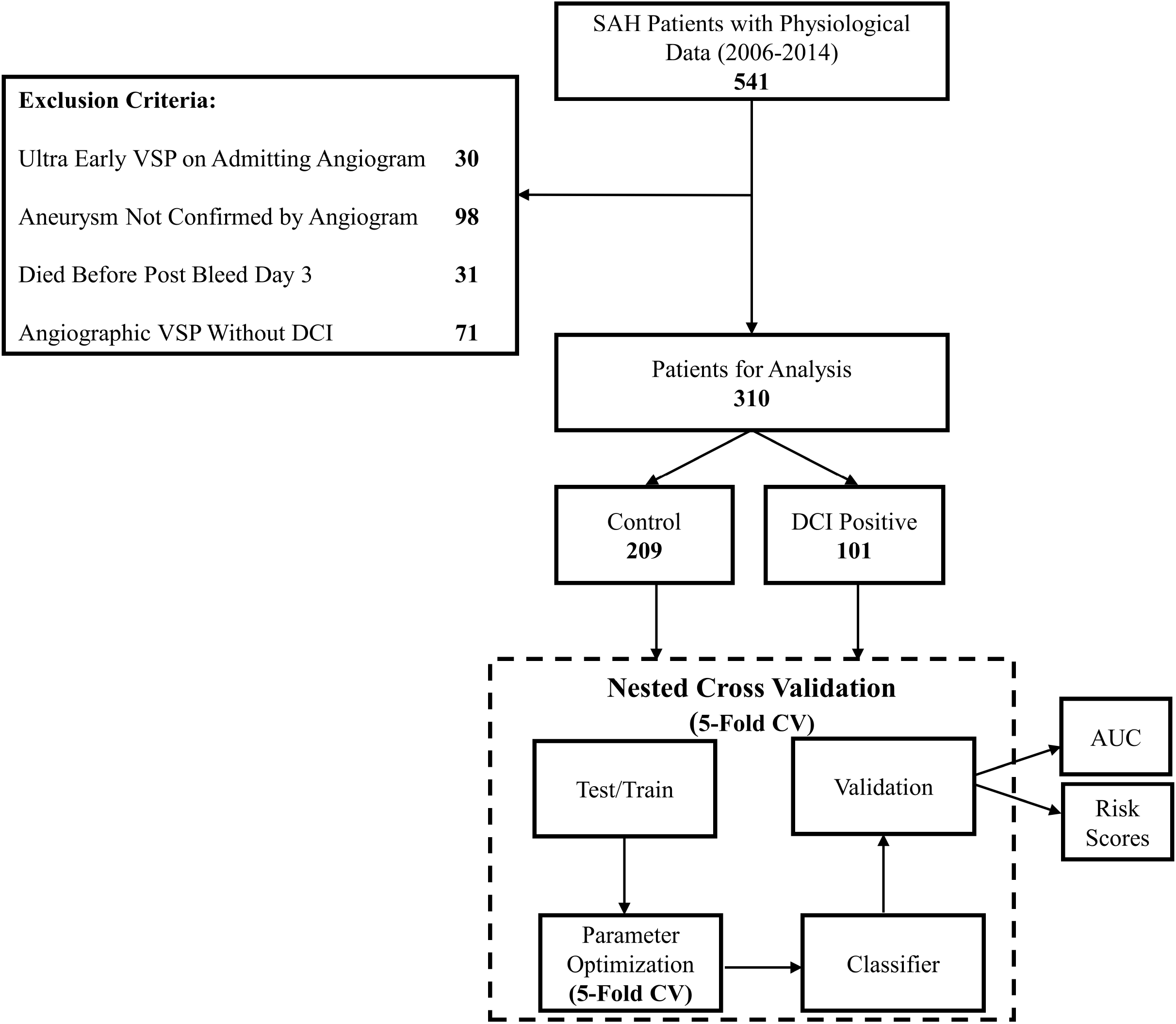
Overview of the approach.

### Multilevel Linear Regression Analysis

At DCI anchor, hourly averages of mean HR, max cross-correlation of SPO2-HR and standard deviation of HR, RR and DBP were statistically different (p<0.001) (**Figure 2**). This statistical approach demonstrated additional significant between-group differences scattered across vitals and time-points. However, there was notable overlap in the distribution of data, indicating the need for more sophisticated methods to classify individual patients.

**Figure 2:**
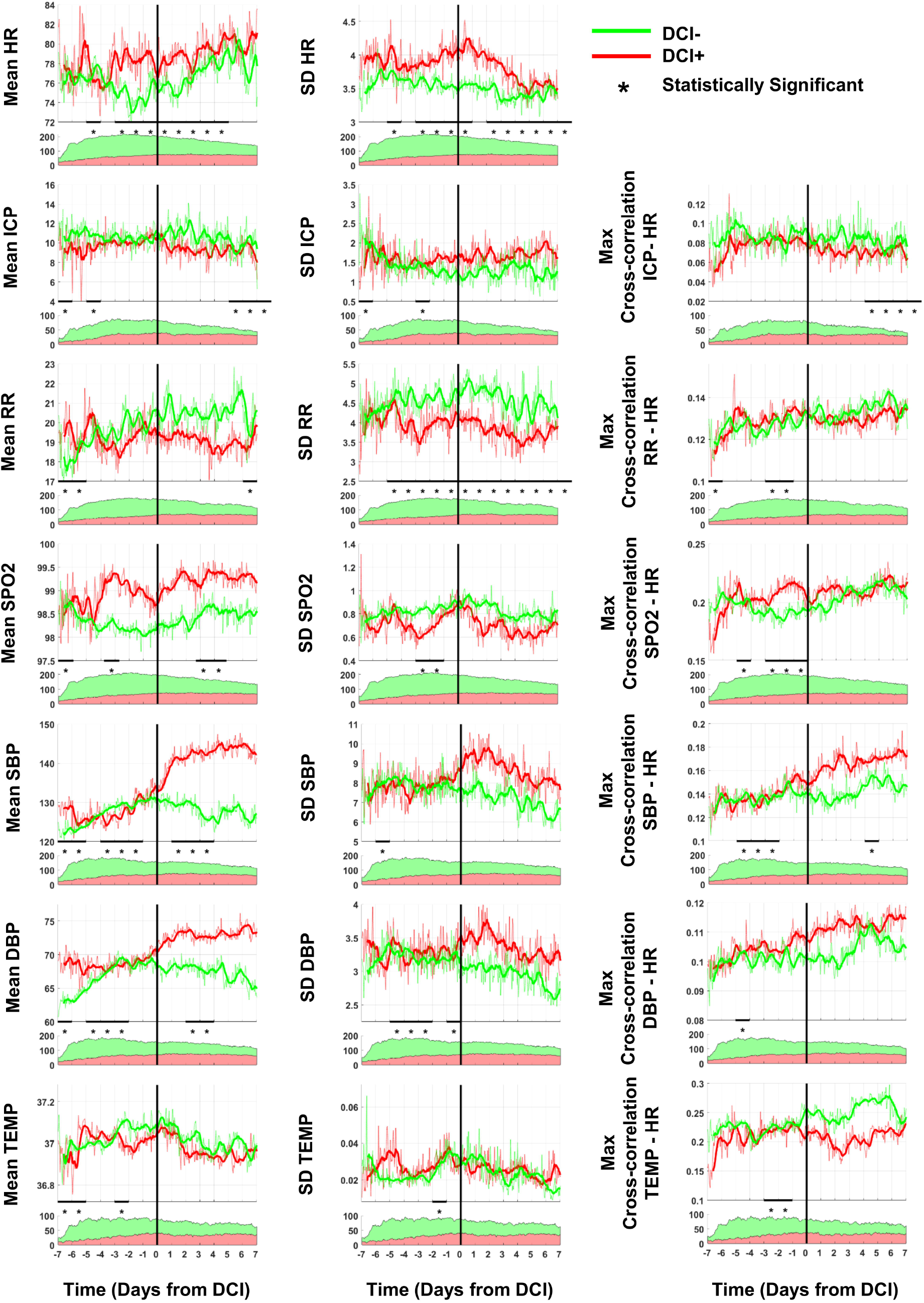
Comparison of Vital Sign Time Series. Multilevel linear regression is used to demonstrate significant differences (*) between DCI+ (red) and DCI– (green) patient vitals. The 20 variables included are mean and standard deviation (SD) of heart rate (HR), intracranial pressure (ICP), respiratory rate (RR), oxygen saturation (SPO2), systolic blood pressure (SBP), diastolic blood pressure (DBP), temperature (TEMP) and maximum cross-correlation of HR with each vital sign. Black vertical line represents DCI anchor. Time axis is a count of days before and after anchor. Number of patients with available data is plotted below each variable, across the same time axis.

### Predictive Models (Best Performing One Used to Generate the Hourly Risk Score)

All classifiers trained on cumulative vitals features up to the DCI anchor performed better than characteristic features alone (**Figure 3A**). Maximal classification was achieved using all features (cumulative vitals + characteristic features), producing AU-ROCs: LR = 0.78 [0.71-0.80], SL = 0.76 [0.75-0.81], SK = 0.76 [0.70-0.78], RF = 0.81 [0.75-0.82] and EC = 0.79 [0.75-0.80]. The performance of classifiers trained with all data up to the anchor produced AU-ROCs: LR = 0.67 [0.66-0.69], SL = 0.65 [0.63-0.67], SK = 0.72 [0.67-0.74], RF = 0.74 [0.74-0.76] and EC = 0.75 [0.70-0.78] (**Table 2**). Classifier weights (LR, SL, RF) indicating the discriminative power of the features in separating the two classes are shown in **Appendix F**.

**Table 2.** AU-ROCs for 5 classifiers at all time-points.

| Time from DCI | LR | SL | SK | RF | EC |
| --- | --- | --- | --- | --- | --- |
| -6 | 0.71 [0.65-0.76] | 0.65 [0.63-0.77] | 0.66 [0.66-0.82] | 0.71 [0.64-0.79] | 0.70 [0.68-0.80] |
| -5.5 | 0.72 [0.62-0.72] | 0.63 [0.62-0.75] | 0.65 [0.65-0.73] | 0.68 [0.68-0.69] | 0.74 [0.65-0.75] |
| -5 | 0.57 [0.52-0.57] | 0.64 [0.5 -0.68] | 0.50 [0.49-0.65] | 0.72 [0.56-0.77] | 0.70 [0.57-0.71] |
| -4.5 | 0.64 [0.44-0.78] | 0.64 [0.58-0.71] | 0.64 [0.56-0.71] | 0.75 [0.74-0.78] | 0.72 [0.65-0.81] |
| -4 | 0.55 [0.51-0.67] | 0.56 [0.52-0.71] | 0.62 [0.55-0.66] | 0.68 [0.64-0.72] | 0.65 [0.59-0.78] |
| -3.5 | 0.67 [0.54-0.79] | 0.65 [0.57-0.65] | 0.63 [0.61-0.68] | 0.78 [0.73-0.79] | 0.74 [0.61-0.76] |
| -3 | 0.67 [0.54-0.70] | 0.53 [0.51-0.57] | 0.61 [0.59-0.66] | 0.72 [0.71-0.80] | 0.71 [0.65-0.75] |
| -2.5 | 0.64 [0.55-0.70] | 0.58 [0.58-0.64] | 0.63 [0.55-0.67] | 0.76 [0.61-0.78] | 0.70 [0.6 -0.73] |
| -2 | 0.60 [0.58-0.64] | 0.62 [0.6 -0.69] | 0.69 [0.65-0.72] | 0.75 [0.66-0.76] | 0.70 [0.61-0.75] |
| -1.5 | 0.66 [0.61-0.71] | 0.63 [0.6 -0.66] | 0.70 [0.63-0.70] | 0.74 [0.72-0.75] | 0.69 [0.68-0.73] |
| -1 | 0.66 [0.65-0.68] | 0.63 [0.62-0.64] | 0.67 [0.66-0.67] | 0.67 [0.66-0.72] | 0.69 [0.68-0.72] |
| -0.5 | 0.66 [0.66-0.72] | 0.66 [0.64-0.70] | 0.71 [0.66-0.76] | 0.77 [0.69-0.8 ] | 0.74 [0.67-0.8 ] |
| **0** | **0.67 [0.66-0.69]** | **0.65 [0.63-0.67]** | **0.72 [0.67-0.74]** | **0.74 [0.74-0.76]** | **0.75 [0.70-0.78]** |
| 0.5 | 0.67 [0.66-0.71] | 0.67 [0.67-0.69] | 0.74 [0.66-0.74] | 0.74 [0.72-0.79] | 0.75 [0.68-0.77] |
| 1 | 0.66 [0.64-0.73] | 0.65 [0.63-0.72] | 0.66 [0.65-0.78] | 0.74 [0.71-0.80] | 0.68 [0.66-0.77] |
| 1.5 | 0.67 [0.66-0.74] | 0.68 [0.66-0.76] | 0.68 [0.68-0.78] | 0.75 [0.71-0.76] | 0.68 [0.68-0.78] |
| 2 | 0.72 [0.68-0.72] | 0.73 [0.68-0.73] | 0.73 [0.70-0.75] | 0.77 [0.7 -0.78] | 0.76 [0.71-0.78] |
| 2.5 | 0.73 [0.7 -0.74] | 0.75 [0.7 -0.76] | 0.71 [0.70-0.75] | 0.78 [0.74-0.79] | 0.78 [0.71-0.78] |
| 3 | 0.76 [0.69-0.77] | 0.76 [0.69-0.78] | 0.75 [0.70-0.77] | 0.76 [0.76-0.77] | 0.78 [0.71-0.79] |
| 3.5 | 0.73 [0.69-0.78] | 0.76 [0.7 -0.76] | **0.76 [0.70-0.78]** | 0.77 [0.76-0.79] | 0.76 [0.71-0.79] |
| 4 | 0.73 [0.71-0.76] | 0.72 [0.69-0.77] | 0.75 [0.73-0.79] | **0.81 [0.75-0.82]** | 0.77 [0.75-0.79] |
| 4.5 | **0.78 [0.71-0.80 ]** | 0.72 [0.72-0.74] | 0.73 [0.71-0.78] | 0.80 [0.78-0.82] | **0.79 [0.75-0.8 ]** |
| 5 | 0.77 [0.73-0.79] | 0.74 [0.72-0.74] | 0.75 [0.68-0.76] | 0.80 [0.80-0.81] | 0.78 [0.76-0.79] |
| 5.5 | 0.77 [0.74-0.79] | 0.75 [0.75-0.75] | 0.76 [0.75-0.76] | 0.75 [0.74-0.79] | 0.77 [0.76-0.8 ] |
| 6 | 0.77 [0.75-0.8 ] | 0.74 [0.73-0.76] | 0.76 [0.76-0.76] | 0.78 [0.77-0.78] | 0.78 [0.78-0.8 ] |
| 6.5 | 0.77 [0.75-0.81] | 0.76 [0.75-0.78] | 0.74 [0.74-0.79] | 0.76 [0.76-0.78] | 0.78 [0.77-0.82] |
| 7 | 0.77 [0.75-0.83] | **0.76 [0.75-0.81]** | 0.75 [0.74-0.75] | 0.77 [0.75-0.80] | 0.78 [0.77-0.82] |

**Figure 3:**
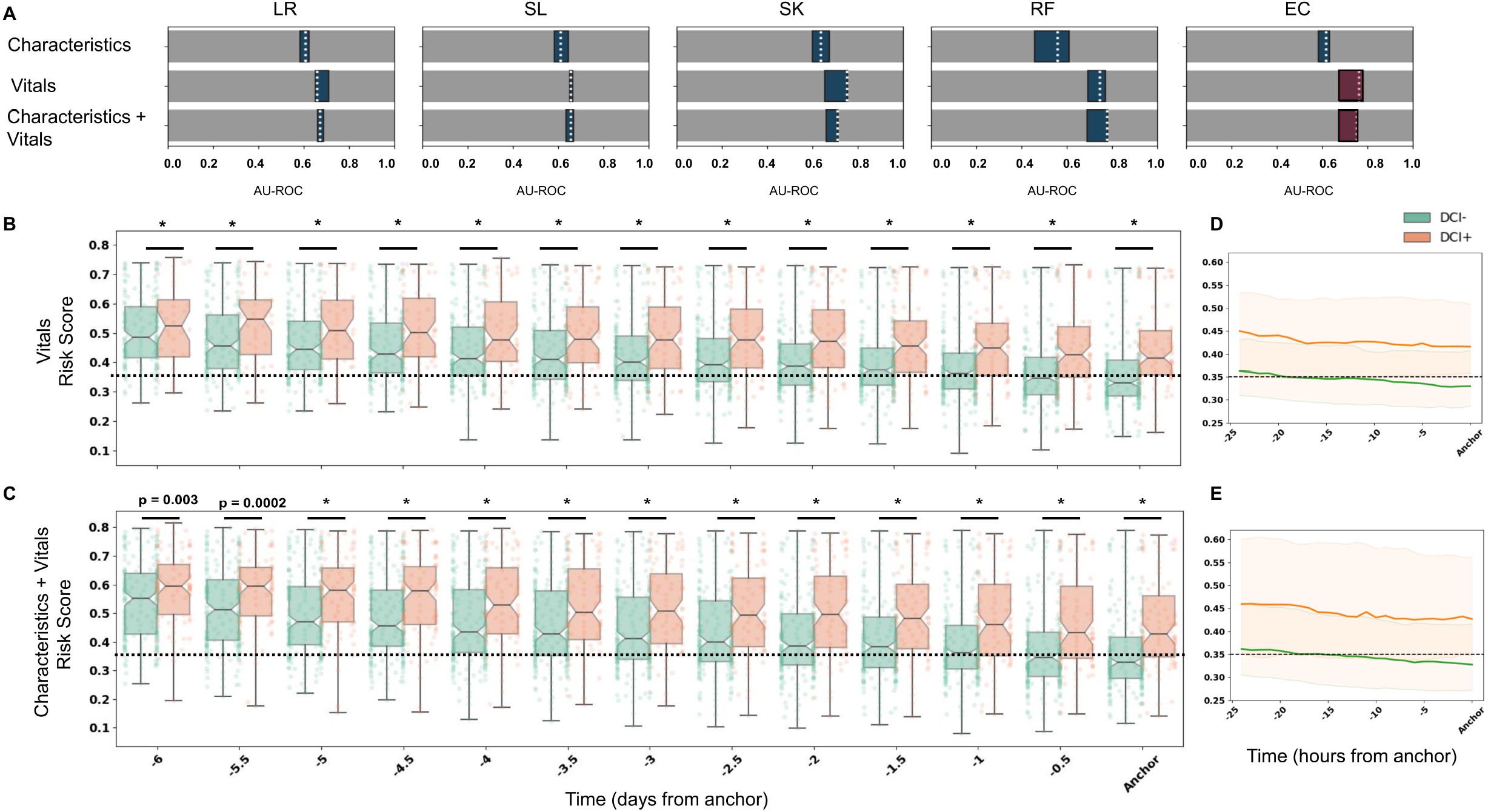
Classifier performance and risk scores. (A) AU-ROCs for five classifiers (LR, SL, SK, RF and EC) trained on characteristic and vitals features at DCI anchor. Blue box indicates the IQR and white dotted lines are the median AU-ROCs. Best performing models are highlighted in red and the risk were generated using these models. Risk scores with based on the models created with (B) vital features alone and (C) characteristics + vitals are shown every 12 hours. The risk scores for DCI+ (red) and DCI- (green) were significantly different at all time-points. Black dotted line represents the optimal threshold (0.35). The median risk score for DCI-patients drops below the threshold 12 hours prior to anchor (D&E).

### Risk Scores

We computed hourly risk scores using the EC classifiers with the highest performance before the DCI anchor, with (**Figure 3C**) and without characteristic features (**Figure 3B**). The optimal threshold (where true positive rate is high and the false positive rate is low) for EC was 0.35. DCI+ and DCI-risk scores are initially high. Over time DCI-patients’ scores drop below the threshold, while DCI+ patients’ scores remain above the threshold (**Figure 3D-E**). With a threshold of 0.35, our models predicted 72.6 % (with) and 74.7 % (without) of all DCI events 12 hours before typical clinical detection. In other words, about 3 true alerts for every 2 false alerts were generated. This threshold can be adjusted to reduce the false negative rate, however, this would be at the expense of a higher false positive rate.

## Discussion

A cohort of 310 patients with aneurysmal SAH, prospectively collected and adjudicated for DCI between June 2006 and December 2014, were retrospectively analyzed and included in the final study. We generated hourly risk scores for the detection of DCI using machine learning models trained on physiologic time series data. DCI positive patients received high risk scores at 12 hours before diagnosis, and patients who did not develop DCI received low risk scores. There is an abundance of routinely collected physiological data generated in the NICU. To our knowledge, this is the first study in humans to apply a data-driven automated approach using vital sign time series data to produce hourly classifications of DCI after SAH.

The current practice for DCI diagnosis is heavily reliant on availability of a useful exam in already neurologically injured patients (20% of SAH patients are comatose)^31^ as well as the attention and expertise of caregivers in the busy and diurnal hospital setting.^32^ An automated, continuous monitoring tool has the potential to provide new insight, and conveniently, our risk scores are derived from routinely collected vitals data (HR, ICP, RR, SPO2, SBP, DBP and TEMP). In this study we calculated cross-correlations to featurize the relationships between HR and the other vital signs. The cross-correlation method should support good generalizability across institutions; the statistic describes co-trending of vital signs rather than absolute values, which would be much more vulnerable to institutional protocol variation.^33^

Notably, a similar strategy was used to identify a preclinical signal in neonates who develop necrotizing enterocolitis or late-onset septicemia, diseases that likely share the cumulative deleterious inflammatory pathway in DCI.^33^ While the exact pathophysiology of DCI is multifactorial, vagal (parasympathetic) dysfunction ^34,35^ and inflammation ^36-39^ are recognized in the pathway, and intricately related to each other (the inflammatory reflex).^40-45^ It was therefore reasonable to expect that evidence of this mechanism could be accessed through routinely collected physiologic data.

Initial efforts towards automated DCI classification showed that machine learning classifiers trained on simple summary statistics of passively-obtained vitals and lab information significantly out-performed classifiers trained on resource-intensive information such as TCDs and nursing exams.^46^ Later work combined time series analysis of vitals and more sophisticated machine learning methods ^10,11^ to discover hidden signals of patient state within continuous physiologic data. Each of these studies incorporated information from just the first few days post-bleed, and boosted prediction to 0.77–0.78 AU-ROC, surpassing the current gold standard for DCI prediction (mFS) at 0.54–0.58 AU-ROC. The success of these machine learning models supports the existence of a useful physiologic signal for the classification of patients with DCI, however these are still prediction models that do not provide an estimation of disease onset time. We were able to see similarly high AU-ROCs in our models trained on just the first few days of data, but as we included data sequentially closer to DCI onset, our model performance continued to increase. The risk scores also demonstrate the advantage of continuously adding data. Both DCI+ and DCI-patients begin with high risk scores, and as the patient state is updated the risk scores diverge over time.

There are several limitations to this study. First, since the range of time to DCI onset after SAH is large, when we align patients by individual diagnosis, the amount of data before and after the anchor is inconsistent across patients. We tolerate this variation because alignment by the outcome event should maximize the information in the physiologic signal leading up to it, and our priority is to create models that can best capture this signal. Another limitation concerns generalizability. We applied nested fivefold cross-validation to avoid overfitting the data, but testing is needed on additional cohorts to sufficiently validate this model for generalizability. In future work we plan to apply these methods and include an external validation protocol with another institution.

## Conclusion

DCI after SAH contributes to morbidity and mortality of NICU patients. Early detection of DCI before permanent secondary brain injury is a clinical goal in SAH patient management, in an effort to improve outcomes. We generated hourly risk scores for the detection of DCI using machine learning models trained on physiologic time series data and subject characteristics. DCI+ patients received high risk scores (above threshold) and DCI– patients received low risk scores (below threshold) before DCI anchor. Future efforts will focus on how to further improve the precision of DCI prediction, validating on external cohorts, and translating these metrics to real-time bedside displays to test their impact on outcomes in future clinical trials.

## Online Supplement

***Appendix A:*** Clinical Management of Subarachnoid Hemorrhage Patients

***Appendix B:*** Anchoring

***Appendix C***: Multilevel Linear Regression

***Appendix D***: Machine Learning

***Appendix E***: Nested Cross-Validation

***Appendix F***: Classifier Weights

